# Severity-stratified and longitudinal analysis of VWF/ADAMTS13 imbalance, altered fibrin crosslinking and inhibition of fibrinolysis as contributors to COVID-19 coagulopathy

**DOI:** 10.1101/2020.08.18.20159608

**Authors:** Kieron South, Lucy Roberts, Lucy Morris, Elizabeth R. Mann, Madhvi Menon, Sean Blandin Knight, Joanne E. Konkel, Andrew Ustianowski, Nawar Diar Bakerly, Paul Dark, Angela Simpson, Timothy Felton, Alexander Horsley, CIRCO, Tracy Hussell, John R. Grainger, Craig J. Smith, Stuart M. Allan

**Affiliations:** Lydia Becker Institute of Immunology and Inflammation, School of Biological Sciences, Faculty of Biology, Medicine and Health, University of Manchester, Manchester Academic Health Science Centre, Room 2.16, Core Technology Facility, 46 Grafton Street, Manchester, M13 9PL, UK; Division of Neuroscience and Experimental Psychology, School of Biological Sciences, Faculty of Biology, Medicine and Health, The University of Manchester, Manchester Academic Health Science Centre, AV Hill Building, Manchester, M13 9PT; Division of Infection, Immunity and Respiratory Medicine, School of Biological Sciences, Faculty of Biology, Medicine and Health, The University of Manchester, Manchester Academic Health Science Centre, AV Hill Building, Manchester, M13 9PL; Maternal and Fetal Health Centre, Division of Developmental Biology, School of Medical Sciences, Faculty of Biology, Medicine and Health, The University of Manchester, 5th Floor St. Mary’s Hospital, Oxford Road, Manchester M13 9WL, UK; Regional Infectious Diseases Unit, North Manchester General Hospital, Manchester, UK; Department of Respiratory Medicine, Salford Royal NHS Foundation Trust, Stott Lane, M6 8HD, UK; Intensive Care Department, Salford Royal NHS Foundation Trust, Stott Lane, M6 8HD, UK; Division of Infection, Immunity and Respiratory Medicine, Manchester NIHR BRC, Education and Research Centre, Wythenshawe Hospital, UK; Division of Cardiovascular Sciences, Faculty of Biology, Medicine and Health, The University of Manchester, Manchester Academic Health Science Centre, Manchester, M13 9PT; Manchester Centre for Clinical Neurosciences, Manchester Academic Health Science Centre, Salford Royal NHS Foundation Trust, Salford M6 8HD

## Abstract

**Background:** Early clinical reports have suggested that the prevalence of thrombotic complications in the pathogenesis of COVID-19 may be as high as 30% in intensive care unit (ICU)-admitted patients and could be a major factor contributing to mortality. However, mechanisms underlying COVID-19-associated thrombo-coagulopathy, and its impact on patient morbidity and mortality, are still poorly understood.

**Methods:** We performed a comprehensive analysis of coagulation and thromboinflammatory factors in plasma from COVID-19 patients with varying degrees of disease severity. Furthermore, we assessed the functional impact of these factors on clot formation and clot lysis.

**Results:** Across all COVID-19 disease severities (mild, moderate and severe) we observed a significant increase (6-fold) in the concentration of ultra-large von Willebrand factor (UL-VWF) multimers compared to healthy controls. This is likely the result of an interleukin (IL)-6 driven imbalance of VWF and the regulatory protease ADAMTS13 (a disintegrin and metalloproteinase with thrombospondin type 1 motifs, member 13). Upregulation of this key pro-coagulant pathway may also be influenced by the observed increase (~6-fold) in plasma α-defensins, a consequence of increased numbers of neutrophils and neutrophil activation. Markers of endothelial, platelet and leukocyte activation were accompanied by increased plasma concentrations of Factor XIII (FXIII) and plasminogen activator inhibitor (PAI)-1. In patients with high FXIII we observed alteration of the fibrin network structure in *in vitro* assays of clot formation, which coupled with increased PAI-1, prolonged the time to clot lysis by the t-PA/plasmin fibrinolytic pathway by 52% across all COVID-19 patients (n=23).

**Conclusions:** We show that an imbalance in the VWF/ADAMTS13 axis causing increased VWF reactivity may contribute to the formation of platelet-rich thrombi in the pulmonary vasculature of COVID-19 patients. Through immune and inflammatory responses, COVID-19 also alters the balance of factors involved in fibrin generation and fibrinolysis which accounts for the persistent fibrin deposition previously observed in post-mortem lung tissue.

**What is new?:**

- In all COVID-19 patients, even mild cases, UL-VWF is present in plasma due to the alteration of VWF and ADAMTS13 concentrations, likely driven by increased IL-6 and α-defensins.
- Increased plasma FXIII alters fibrin structure and enhances incorporation of VWF into fibrin clusters.
- Defective fibrin structure, coupled with increased plasma PAI-1 and α2-antiplasmin, inhibits fibrinolysis by t-PA/plasmin.

**What are the clinical implications?:**

- Prophylactic anticoagulation and management of thrombotic complications in COVID-19 patients are ongoing challenges requiring a better understanding of the coagulopathic mechanisms involved.
- We have identified FXIII and VWF as potential therapeutic targets for treating fibrin formation defects in COVID-19 patients.
- We have identified a multifaceted fibrinolytic resistance in COVID-19 patient plasma with potential implications in the treatment of secondary thrombotic events such as acute ischaemic stroke or massive pulmonary embolism.

## Introduction

The severe acute respiratory syndrome coronavirus (SARS-CoV)-2 pandemic, since its origin in December 2019, has affected more than 20 million individuals worldwide resulting in almost 750,000 fatalities. The diseased state induced by the virus, coronavirus disease 2019 (COVID-19), manifests as a broad clinical spectrum ranging from asymptomatic individuals, through to a mild flu-like illness and, in a small proportion of people, a severe hyper-inflammatory state causing acute respiratory distress syndrome (ARDS), multiple organ failure and death. Early clinical and post-mortem observations also indicated the existence of a hyper-coagulative state, seemingly distinct from sepsis-induced disseminated intravascular coagulation (DIC), which was widespread amongst critically-ill patients^1, 2^.

Subsequent studies have estimated an incidence of thrombotic events (mainly pulmonary embolism and venous thromboembolism) in the range of 15-30%, but as high as 69%, of intensive care unit (ICU)-admitted patients^3-6^. Moreover, thrombotic complications have been reported in an estimated 6% of patients across general wards, all of whom had received prophylactic anticoagulants^6^. Potential manifestations of this hyper-coagulative state are acute limb ischaemia^7^, deep vein thrombosis^8^, pulmonary embolism and ischaemic stroke^8-11^. Early estimates of ischaemic stroke prevalence amongst COVID-19 admissions range from 0.9% to 5% amongst severe cases^11, 12^. Interestingly, one small case series identified an apparent increased risk in young adults, many of whom presented with stroke and asymptomatic SARS-CoV-2 infection^12^. This may suggest that thrombotic complications are not restricted to those at risk of more severe disease progression. Definitive evidence, from large prospective studies, for a causal relationship between COVID-19 and stroke is still required^13^.

There has been much discussion postulating possible mechanisms underlying the COVID-19 associated coagulopathy and the idea that immunothrombosis, or thromboinflammation, is driving the pro-coagulant state is beginning to gain prominence. Recent studies have identified altered platelet reactivity (and increased platelet-leukocyte interactions)^14^, release of neutrophil extracellular traps (NETs)^15^ and endothelial activation^16^ in COVID-19 patients. Several studies have identified an increase in plasma VWF associated with COVID-19^16-18^, however, these have concentrated on those with severe disease (ICU versus non-ICU) and have not included concurrent longitudinal analyses of both VWF and other thromboinflammatory markers. As such, the mechanisms underlying the presence of VWF and how this relates to patient morbidity, mortality or recovery remains unclear.

The extremely high and persistent D-dimer levels observed in some COVID-19 patients are an indicator of poor prognosis. They indicate ongoing fibrin formation and fibrinolysis but may also suggest that, while still functional, the fibrinolytic system is overwhelmed^19^. Whether this is due to enhanced fibrinolytic resistance or direct inhibition of pro-fibrinolytic factors is unknown. The former may result from a local increase in Factor XIII which, through its fibrin crosslinking activity, stabilises fibrin clots. This is due to both a direct influence of fibrin structure^20^ and the incorporation of α2-antiplasmin into the fibrin network^21^. Direct inhibition of fibrinolysis is mediated by the soluble factors plasminogen activator inhibitor-1 (PAI-1) and thrombin-activatable fibrinolysis inhibitor (TAFI), both of which interfere with the normal fibrinolytic activity of tissue plasminogen activator (t-PA). A combination of these processes might be expected to produce persistent fibrin deposits that are resistant to fibrinolysis, as is suggested by COVID-19 lung pathology.

The aim of this study is to identify potential haemostatic deficits, and underlying thromboinflammatory mechanisms, in COVID-19 patient plasma using a combination of clinical measurements and functional coagulation assays. The relationship between these parameters and their significance in disease progression and outcome have been addressed by the application of disease severity stratification and longitudinal sampling.

## Methods

### Study Design

Between 29^th^ March and 7^th^ May 2020, patients admitted with suspected COVID-19 were recruited across hospitals in the Greater Manchester area. Peripheral blood samples were collected at Manchester University Foundation Trust (MFT), Salford Royal NHS Foundation Trust (SRFT) and Pennine Acute NHS Trust (PAT) under the framework of the Manchester Allergy, Respiratory and Thoracic Surgery (ManARTS) Biobank (study no M2020-88) for MFT or the Northern Care Alliance Research Collection (NCARC) tissue biobank (study no NCA-009) for SRFT and PAT (REC reference 15/NW/0409 for ManARTS and 18/WA/0368 for NCARC). Informed consent was obtained for each patient and clinical information was extracted from written/electronic medical records. Patients were eligible for the study if they tested positive for SARS-CoV-2 by reverse-transcriptase-polymerase-chain-reaction (RT-PCR) of nasopharyngeal swaps, or, in the absence of a positive RT-PCR test, by high clinical suspicion of COVID-19 with characteristic radiological findings. Patients were excluded on the basis of a negative RT-PCR combined with indeterminate radiological findings, if an alternate diagnosis was reached or if the patient had a confounding acute illness.

Severity stratification of recruited patients was based on the degree of respiratory failure. Mild cases were characterised by a supplemental oxygen requirement of <3l or <28% and management in a ward-based environment. Moderate cases required <10l or <60% supplemental oxygen and management in a ward-based environment. Those determined to have severe disease required >10l or >60% supplemental oxygen, were managed in ICU and/or required invasive ventilation. When severity levels changed during admission, patients were stratified by their highest disease severity score. All cross-sectional comparisons between the groups were performed using the first available sample, collected as soon as possible after admission. Longitudinal samples for a sub-set of severe patients were collected at 1-2 day intervals.

### Healthy controls

Four healthy control plasma samples were obtained from Manchester University and NHS Trust front line staff at the time of collecting patient plasma. These individuals were asymptomatic and assumed COVID-19 negative but were not tested. A further 16 healthy controls samples, pre-dating the COVID-19 pandemic, were obtained as part of a previous study and were included in these analyses to ensure age/sex matching of controls to COVID-19 patient samples. Of these, 4 were receiving aspirin but none were otherwise anti-coagulated.

### Plasma preparation

Whole blood, collected using BD Vacutainers containing 3.2% trisodium citrate, was diluted 2-fold in phosphate buffered saline (PBS) and fractionated by density gradient centrifugation. Platelet rich plasma was removed and centrifuged further to deplete platelets. Platelet poor plasma was stored at -80°C for analysis. Total plasma protein concentration was quantified by BCA assay (Pierce) following manufacturer’s protocol.

### Multiplex assays

An initial screen of coagulation factors was performed on a small cohort of healthy control (n=4) and COVID-19 patient plasma (n=12) using Procartaplex (Thermo-Fisher) mix and match coagulation panels (Factors X, XI, XII, V, VII, VIII, Protein C and Protein S). All were performed following manufacturer’s instructions and analysed on a Bio-Plex 200 Luminex analyser (Bio-Rad).

All subsequent analyses were performed on the full cohort of healthy control (n=20) and COVID-19 patient plasma (n=23) using the following LEGENDplex panels. Human inflammation 13-plex panel (IL-1β, IFN-α2, IFN-γ, TNF-α, MCP-1, IL-6, IL-8, IL-10, IL-12p70, IL-17A, IL-18, IL-23 and IL-33) human thrombosis custom 7-plex panel (P-selectin, D-dimer, PSGL-1, t-PA, sCD40L, tissue factor and Factor IX) and human fibrinolysis 5-plex panel (fibrinogen, plasminogen, antithrombin, prothrombin and Factor XIII). All were performed using the manufacturer’s protocol and suggested dilutions. Analysis was performed using a BD FACSVerse flow cytometer (BD Biosciences) and LEGENDplex data analysis software version 8.0.

### Quantification of Thromboinflammatory Markers in Plasma by ELISA

Plasma VWF and ADAMTS13 antigen levels were determined by in-house ELISA as described previously^22^, with intra- and inter- assay coefficients of variation of 5 and 8% respectively for VWF and 8 and 12% respectively for ADAMTS13^23^.

The presence of UL-VWF in plasma was determined by an ELISA-based collagen binding assay^23^. Results are presented as both VWF:CBA (%), in which the signal is normalised to that of healthy controls, and the ratio of VWF:CBA/Ag, in which the extent of collagen binding is normalised by the total VWF antigen determined by ELISA.

Commercial ELISA kits were used, following manufacturer’s protocols, to quantify α -defensins (human HNP (1-3) ELISA, Hycult Biotech), α2-antiplasmin (human Serpin F2 ELISA, Invitrogen), TAFI (human carboxypeptidase B2/CPB2 ELISA, Invitrogen) and PAI-1 (Invitrogen).

### Turbidity assay of fibrin formation

Sample plasma was warmed to 37°C and diluted 2-fold in Hank’s Balanced Salt Solution (HBSS) supplemented with 20 mM CaCl_2_ in clear, flat-bottomed 96-well plates in a final volume of 50 μl. The assay was initiated by the addition of 5 nM (0.5 U/ml) human thrombin (Sigma-Aldrich) at which point the samples were mixed for 10 seconds at 700 rpm in the heated chamber of a FluoSTAR Omega microplate reader (BMG Labtech) pre-heated to 37°C. The absorbance at 405 nm was measured at 20 second intervals for a total of 30 minutes.

### Confocal microscopy of fibrin structure

Plasma from 3 healthy controls and 3 patients in each COVID-19 subgroup was diluted as above, in a total volume of 200 μl in 12-well chamber slides. These preparations were supplemented with 150 μg/ml AlexaFluor594 labelled fibrinogen (Invitrogen) and 10 μg/ml AlexaFluor647 conjugated α-VWF antibody (Abcam) and pre-warmed to 37°C. Fibrin polymerisation was initiated by the addition of 5 nM (0.5 U/ml) human thrombin and left to occur at 37°C for 1 hour. Images were acquired on a Leica SP8 inverted confocal microscope equipped with a heated chamber maintained at 37°C. A 10×/0.40 APO dry objective and zoom factor of 0.75 were used to acquire images of 1550 μm × 1550 μm. HyD hybrid detectors were used to detect fibrinogen-594 and VWF-647 with emission gates of 600-642 nm and 691-787 nm, respectively. Images were analysed in ImageJ.

### Fibrinolysis assay

Plasma, diluted as above, was pre-warmed to 37°C. The assay was initiated by the addition of 10 nM (1 U/ml) human thrombin and 25 ng/ml rt-PA (Alteplase, Hoffmann La Roche) at which point the samples were placed in the heated chamber of a FluoSTAR Omega plate reader. Absorbance at 405 nm was measured every 30 s for 1 h with continuous double orbital shaking between measurements.

## Results

### CIRCO patient characteristics

A total of 23 eligible patients, with sufficient observations to allow stratification of disease severity, were recruited with a median time from patient-reported symptom onset to hospital admission of 8 days. The median age was 58 years and 69.6% were male compared with a median age of 67 years and 43.8% male in the healthy control group. The majority of patients (69.6%) tested positive for SARS-CoV-2 by RT-PCR and the remaining 30.4%, with negative RT-PCR, had moderate or severe radiological findings. Frequent co-morbidities amongst patients were diabetes, hypertension (HTN), ischaemic heart disease (IHD), chronic obstructive pulmonary disease (COPD) and asthma (**Table 1**). A total of 82.4% of patients received thromboprophylaxis and 10% received a treatment dose of anti-coagulants. The only complications observed were acute kidney injury (AKI) in 2 moderate patients and pulmonary embolism (PE) in 1 moderate patient (**Table 1**). Death occurred in 1 mild patient (aged 84) and 2 severe patients (aged 74 and 81).

**Table 1.** Clinical characteristics. Data are median (IQR). n (%) or n/N (%), where N is the total number with available data. Numbers in red are the lower detection limit of the assay. ND, not determined in healthy controls. N/A, not applicable to healthy controls.

|  | Healthy controls | All COVID-19 patients | Mild COVID-19 | Moderate COVID-19 | Severe COVID-19 |
| --- | --- | --- | --- | --- | --- |
| **n** | 20 | 23 | 7 | 9 | 7 |
| **Age (Years) [N]** | 67 (53-73.8) [16] | 58 (51-73) | 58 (35-73) | 59 (44.5-74) | 58 (51-74) |
| **Sex (Male)** | 7/16 (43.8 %) | 16 (69.6%) | 4 (57.1%) | 7 (77.8%) | 5 (71.4%) |
| **BMI (kg/m<sup>2</sup>) [N]** | ND | 27.8 (24.7-31) [16] | 27.3 (26.2-31.6) [5] | 28.3 (24.6-31.2) [7] | 27 (23.1-31.8) [4] |
| **Co-morbidity** |  |  |  |  |  |
| Diabetes | Type 2 1/16 (6.3%) | Type 2 4/18 (22.2%) | Type 2 1/6 (16.7%) | Type 2 1/6 (16.7%) | Type 2 2/6 (33.3%) |
| COPD | 0/16 (0%) | 4/19 (21.1%) | 2/6 (33.3%) | 1/7 (14.3%) | 1/6 (16.7%) |
| Asthma | 0/16 (0%) | 1/18 (5.6%) | 0/6 (0%) | 0/6 (0%) | 1/6 (16.7%) |
| IHD | CAD 1/16 (6.3%) | Angina 2/18 (11.1%) | 0/6 (0%) | Angina 1/6 (16.7%) | Angina 1/6 (16.7%) |
| HTN | 4/16 (25%) | 6/18 (33.3%) | 1/6 (16.7%) | 4/6 (66.7%) | 1/6 (16.7%) |
| **Illness onset to sample (days) [N]** | N/A | 8 (4.5-10.8) [20] | 7 (4-10.3) [6] | 7 (4-10) [7] | 9 (7-15) |
| **Presentation** |  |  |  |  |  |
| Respiratory rate (BPM) [N] | ND | 20 (17.5-23) [21] | 20 (19-24) | 18 (17.3-19.8) [8] | 21 (16.8-22.5) [6] |
| O<sub>2</sub> Sats (%) [N] | ND | 96 (91-96.5) [21] | 96 (94-97) | 96 (92.3-97.5) [8] | 90 (86.7-96) [6] |
| Temperature (°C) [N] | ND | 37.7 (37.1-38.4) [21] | 37.4 (36.5-38.1) | 37.7 (37.1-38.8) [8] | 37.8 (37.3-38.7) [6] |
| Systolic blood pressure (mmHg) [N] | ND | 122 (111.5-133.5) [21] | 122 (117-126) | 117.5 (103.5-134.3) [8] | 126.5 (112.5-161) [6] |
| Dyspnoea | N/A | 13/18 (72.2%) | 3 (42.9%) | 4/5 (80%) | 6/6 (100%) |
| Fever | N/A | 14/18 (77.8%) | 5 (71.4%) | 3/5 (60%) | 6/6 (100%) |
| Cough | N/A | 13/18 (72.2%) | 4 (57.1%) | 3/5 (60%) | 6/6 (100%) |
| Myalgia | N/A | 6/18 (33.3%) | 3 (42.9%) | 1/5 (20%) | 2/6 (33.3%) |
| Fatigue | N/A | 2/16 (12.5%) | 0/6 (0%) | 1/4 (25%) | 1/6 (16.7%) |
| **Chest Radiograph Findings** |  |  |  |  |  |
| Bilateral opacification | N/A | 9/12 (75%) | 3/5 (60%) | 3/4 (75%) | 3/3 (100%) |
| Unilateral opacification | N/A | 2/12 (16.7%) | 1/5 (20%) | 1/4 (25%) | 0/3 (0%) |
| No abnormality | N/A | 1/12 (8.3%) | 1/5 (20%) | 0/4 (0%) | 0/3 (0%) |
| **COVID Nasopharyngeal Test (+)** | ND | 16 (69.6%) | 4 (57.1%) | 7 (77.8%) | 5 (71.4%) |
| **Thromboprophylaxis** | Aspirin 4/16 (25%) | 14/17 (82.4%) | 5/6 (83.3%) | 6/6 (100%) | 3/5 (60%) |
| **Antithrombotic Treatment Dose** | 0/16 (0%) | 1/10 (10%) | 1/4 (25%) | 0/3 (0%) | 0/3 (0%) |
| **Admission Full Blood Count [N]** |  |  |  |  |  |
| WCC (x10<sup>9</sup>/l, normal 3.5-10) | 5.5 (4.6-7.8) [16] | 6.6 (5.4-9.8) | 4.7 (4.1-6.9) | 6.6 (6.2-10.4) | 9.4 (6.6-10.1) |
| Lymphocytes(x10<sup>9</sup>/l, normal 0.5-5) | 1.4 (1.2-2.0) [16] | 0.9 (0.8-1.3) | 0.9 (0.78-1.1) | 1.01 (0.9-1.5) | 0.8 (0.8-1.1) |
| Neutrophils(x10<sup>9</sup>/l, normal 2-7.5) | 3.2 (2.3-5.1) [16] | 5.4 (3.7-7.7) | 3.5 (2.6-5.7) | 5.1 (4.6-7.6) | 8.4 (5.4-8.7) |
| Monocytes(x10<sup>9</sup>/l, normal 0.2-0.8) | 0.4 (0.3-0.5) [16] | 0.38 (0.2-0.5) | 0.38 (0.2-0.5) | 0.5 (0.33-0.9) | 0.3 (0.2-0.3) |
| Platelets (x10<sup>9</sup>/l, normal 150-370) | 244.5 (202-294) [16] | 246.5 (190.5-345.8) [20] | 209 (180-263.3) [6] | 338.5 (207.8-472.3) [8] | 264 (149.8-417) [6] |
| **Acute phase responses (IU/L) [N]** |  |  |  |  |  |
| CRP | ND | 116 (23-207) | 54 (16-138) | 68 (19.5-186) | 256 (116-335) |
| ALT | ND | 35 (27-58.5) [9] | 82.5 (82-83) [2] | 59.5 (12.8-92) [4] | 82 (57-176) [3] |
| ALP | ND | 82 (52.5-91) [9] | 36.5 (21-52) [2] | 33.5 (18.8-34.8) [4] | 65 (36-181) [3] |
| Bilirubin | ND | 11 (8-15) [9] | 8 (7-9) [2] | 15 (6.5-19.8) [4] | 11 (9-11) [3] |
| **Complications** |  |  |  |  |  |
| PE | N/A | 1/12 (8.3%) | 0/5 (0%) | 1/4 (25%) | 0/3 (0%) |
| AKI | N/A | 2/12 (16.7%) | 0/5 (0%) | 2/5 (40%) | 0/2 (0%) |
| Mortality | N/A | 3 (13%) | 1 (14.3%) | 0 (0%) | 2 (28.6%) |
| **Clinical D-Dimer (High > 500 ng/ml)** | ND | 4/7 (57%) | 1/1 (100%) | 3/5 (60%) | 0/1 (0%) |
| **Plasma Total Protein (mg/ml)** | 34.2 (31.0-39.3) | 35.1 (29.1-41.1) | 34.7 (30.8-35.1) | 36.5 (24.5-42.9) | 38.7 (24.5-42.6) |
| **Plasma Coagulation Factors** |  |  |  |  |  |
| Plasminogen (µg/ml) | 78 (23.5-192.4) | 80.2 (69.2-107.7) | 80.4 (75.4-92.4) | 80.2 (67.5-112.5) | 72.2 (59.4-110.5) |
| Antithrombin (µg/ml) | 92.3 (30.1-237.6) | 94.8 (69.1-105.6) | 101.4 (82.6-105.6) | 90.3 (65.4-98.3) | 104.3 (73.2-124.9) |
| Prothrombin (µg/ml) | 28.2 (11.0-76.0) | 36.2 (23.9-41.8) | 36.2 (23.1-46.3) | 29.8 (22.8-39.3) | 39.1 (30.1-54.9) |
| Factor IX (ng/ml) | 802 (701-1054) | 1071 (811-1360) | 1002 (425-1118) | 1335 (1078-2091) | 945 (658-2102) |
| **Thrombo-Inflammatory Markers** |  |  |  |  |  |
| IL-12p70 (pg/ml) | 1.3 (1.3-1.31) | 1.3 (1.3-2.19) | 1.3 (1.3-1.44) | 1.3 (1.3-15.7) | 1.69 (1.3-2.77) |
| P-selectin (ng/ml) | 72.5 (55.6-105.7) | 146.8 (78.2-208.6) | 166.6 (91.1-237.4) | 109.5 (40.5-176.1) | 156.2 (88.4-240.0) |
| D-Dimer (ng/ml) | 6.5 (2.2-15.7) | 8.3 (2.8-25.2) | 1.8 (1.3-10.5) | 8.9 (3.9-17.4) | 56.4 (5.8-114.5) |
| sCD40L (ng/ml) | 1.07 (0.72-1.54) | 2.24 (0.69-2.71) | 1.66 (1.28-2.47) | 2.24 (0.35-3.08) | 2.41 (0.61-3.46) |
| PSGL-1 (ng/ml) | 2.24 (1.98-3.12) | 4.04 (2.70-5.06) | 4.04 (3.89-5.09) | 4.1 (2.29-5.29) | 3.07 (1.18-5.06) |

### Peripheral inflammatory markers are increased in COVID-19

The plasma concentrations of the primary anti-viral cytokines IFN-α and TNF-α were not significantly different between COVID-19 patients and healthy controls (**Figure 1**). The peripheral pro-inflammatory response to COVID-19 showed significant increases, compared to healthy controls, for all cytokines except IL-1β and IL-12p70 (**Figure 1 and Table 1**). IL-6, IL-8, IL-10, IL-18, IL-23 and IL-33 were universally increased in all COVID-19 sub-groups compared to healthy controls (**Figure 1**) with IL-6 and IL-8 displaying the largest increase (>40-fold and >4-fold, respectively). Other markers of thromboinflammation (P-selectin, PSGL-1 and sCD40L) were all increased ~2-fold when comparing all COVID-19 samples against healthy controls (**Table 1**), but only P-selectin was statistically significant (p<0.01).

**Figure 1.**
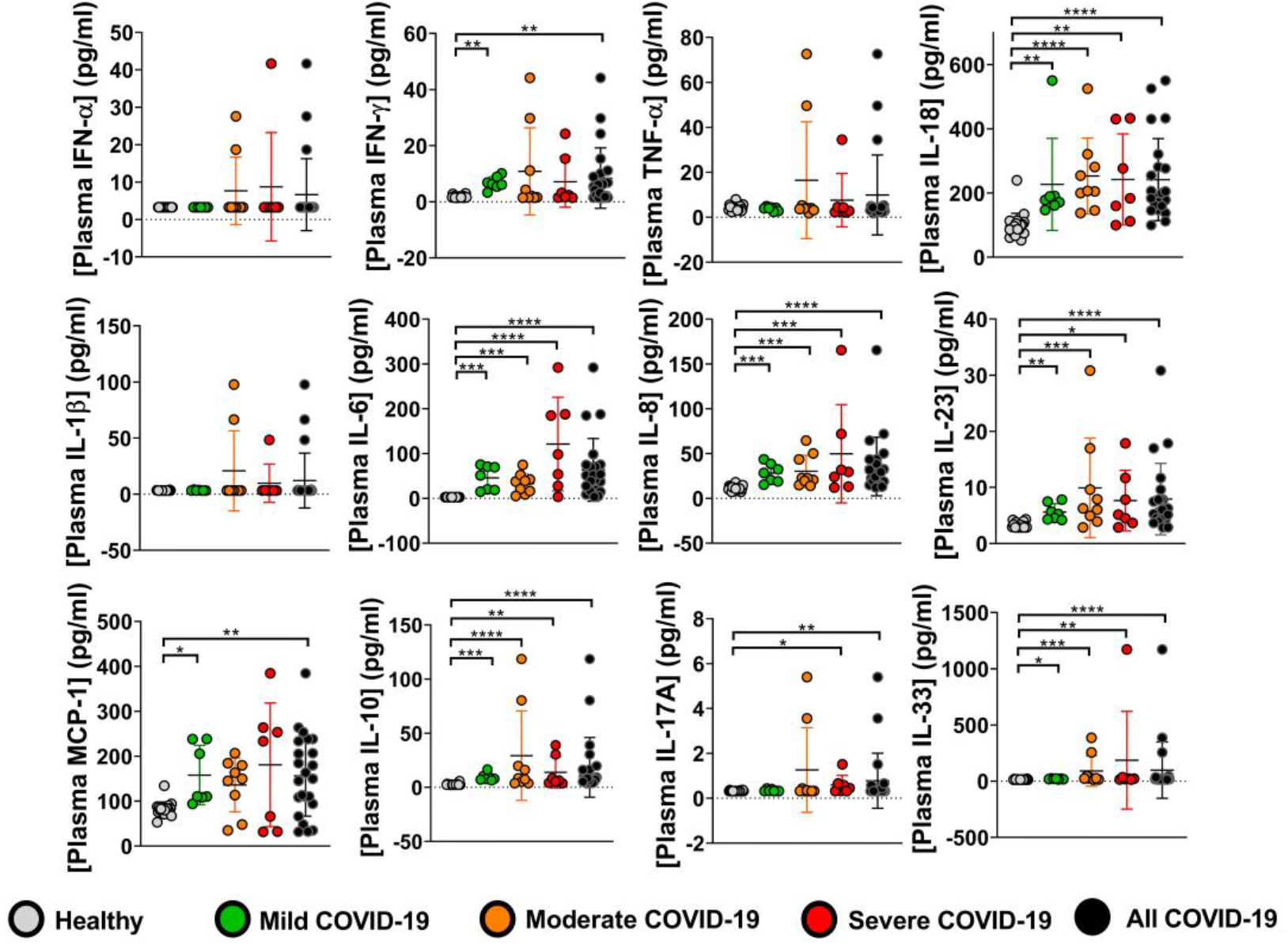
Peripheral inflammatory markers are elevated in plasma from COVID-19 patients. The plasma concentrations of soluble cytokines were determined by LEGENDplex™ assay (human inflammation 13-plex panel) as per manufacturer’s protocol. Each data point represents the mean of duplicate concentration values determined from PE fluorescence intensity. Results are presented as mean ± S.D. for both healthy control (n=20) and COVID-19 patient plasma (n=23). Comparison of the COVID-19 population and healthy controls was performed using a Mann-Whitney test. Comparison of COVID-19 sub-groups with healthy controls was performed using a Kruskal-Wallis test with Dunn’s correction for multiple comparisons (*<0.05, **p<0.01, ***p<0.001 and ****p<0.0001).

Longitudinal sampling was performed on 4 of the severe COVID-19 patients and has suggested some relationships between inflammatory and thrombotic parameters (**Figure S3**). The concentrations of IL-6, IL-8 and MCP-1 all decreased over time in patient 21, peaked at day 3 in patient 20 and increased between day 6 and day 10 in patient 22. In patients 21 and 22 an inverse IL-18 response was observed. A relationship was observed between the inflammatory response (particularly IL-6) and the α-defensin, VWF and PAI-1 responses.

### VWF/ADAMTS13 imbalance is present across all COVID-19 severity groups

An initial Luminex screen of coagulation factor levels in 4 healthy control samples and 12 COVID-19 patient samples identified no significant difference in the PE fluorescence of beads specific to Factors V, VII, VIII, X, XI and XII, Protein S or Protein C (**Figure S1**). Factor IX, antithrombin and prothrombin concentrations were within the normal range with no significant differences between the groups (**Table 1**). Compared to heathy controls, there was a significant increase in D-Dimer concentration (determined by Legendplex) in plasma from the severe COVID-19 group (p=0.004), however, all values were still within the normal range (**Table 1**). The limited number of available results from clinical D-dimer tests identified high concentrations (>500 ng/ml) in some patients (**Table 1**).

A COVID-19 associated increase in the levels of plasma VWF was identified in this initial multiplex screen and was subsequently quantified by ELISA. Healthy controls were all within the normal range (60-190% of 10 μg/ml) with a mean value of 7.5 ± 1.2 μg/ml. This was significantly increased to 25.9 ± 10.8, 19.4 ± 6.6 and 26.1 ± 7.3 μg/ml in the mild, moderate and severe COVID-19 groups respectively (**Figure 2A**). The concentration of ADAMTS13 was also within the normal range (74-142% of 900 ng/ml) for all healthy controls with a mean value of 998 ± 112 ng/ml, but was reduced to 538 ± 163, 582 ± 181 and 589 ± 124 ng/ml in the mild, moderate and severe groups (**Figure 2B**). We also observed a small, but significant, increase (4-6 fold) in neutrophil α-defensins across all COVID-19 sub-groups compared to healthy controls (**Figure 2C**).

**Figure 2.**
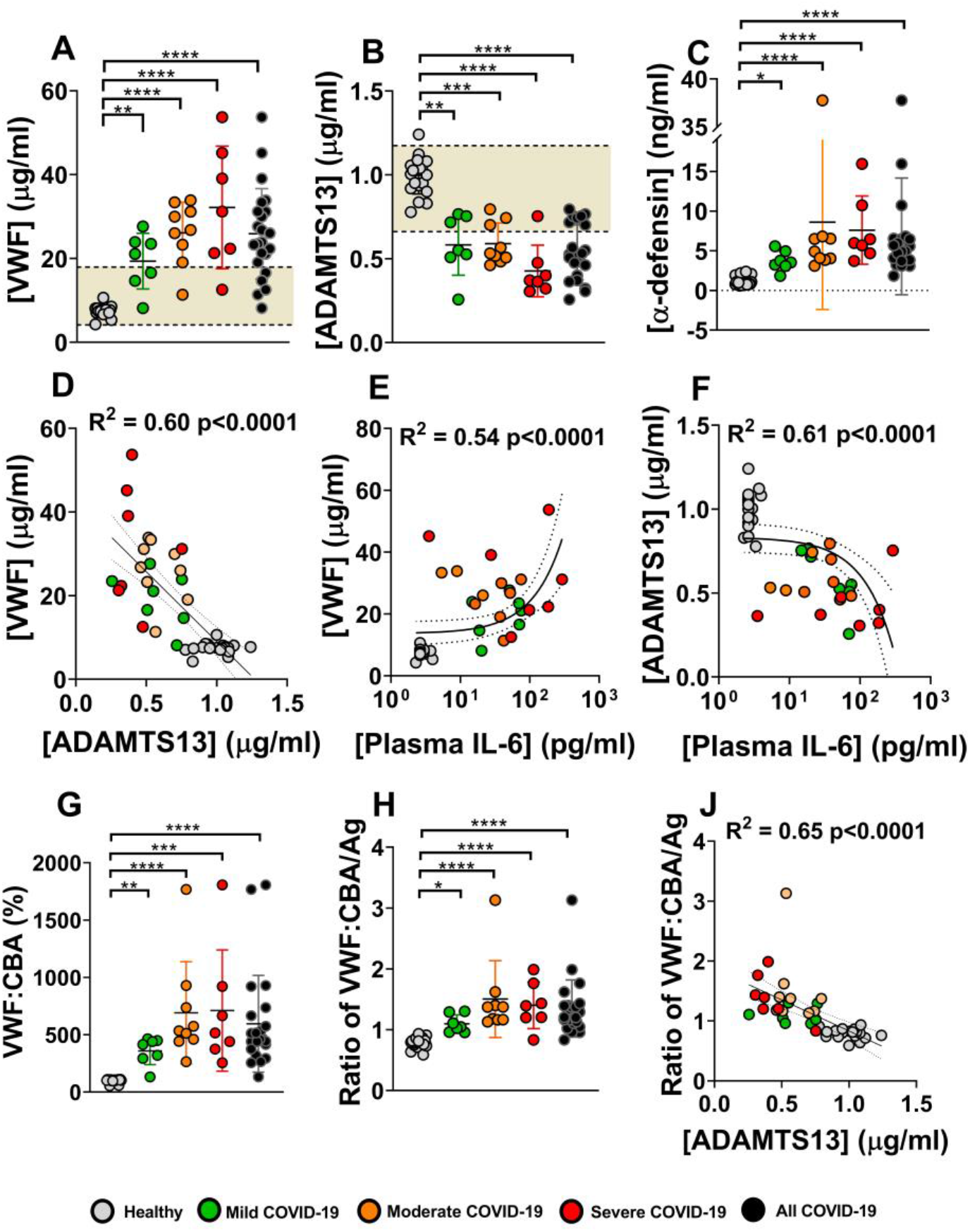
Plasma from COVID-19 patients reveals an IL-6 driven imbalance in the VWF/ADAMTS13 axis, even in patients with mild disease. The plasma concentrations of VWF (**A**) and ADAMTS13 (**B**) were determined by in-house ELISA. The plasma concentrations of α-defensins were determined using a commercial ELISA kit (**C**). Each data point represents the mean of duplicate concentration values determined for each sample. Results are presented as mean ± S.D. for both healthy control (n=20) and COVID-19 patient plasma (n=23). Linear regression (solid line) with 95% confidence intervals (dotted lines) are included to show the relationship between VWF and ADAMTS13 **(D)** and between plasma IL-6 and VWF (**E**) or ADAMTS13 (**F**). In each case Spearman’s rank coefficient correlation tests were performed to determine R^2^ values. The effect of VWF/ADAMTS13 imbalance on the prevalence of high molecular weight (HMW) VWF multimers in plasma was determined by collagen binding assay. The amount of HMW VWF (relative to that in the healthy control group) (**G**) was normalised by total VWF antigen to give the ratio of CBA/Ag (**H**). Linear regression and Spearman’s correlation were performed to show the relationship between HMW VWF and ADAMTS13 **(J)**. Areas shaded in yellow represent the literature values of the normal range. Comparison of the COVID-19 population and healthy controls was performed using a Mann-Whitney test. Comparison of COVID-19 subgroups with healthy controls was performed with a Kruskal-Wallis test with Dunn’s correction for multiple comparisons (*<0.05, **p<0.01 and ****p<0.0001).

A significant inverse relationship between VWF and ADAMTS13 concentration was observed as was a clear segregation of the healthy and patient groups (**Figure 2D**). There was also a significant correlation between VWF and IL-6 concentration (**Figure 2E**) and an inverse relationship between ADAMTS13 and IL-6 (**Figure 2F**). A collagen binding assay was used as a measure of VWF multimeric size and activity with the amount of collagen-bound VWF (VWF:CBA) in each sample normalised to the mean of the healthy control samples (100 ± 15.7 %.). In the mild, moderate and severe COVID-19 groups there was significantly higher VWF activity with mean values of 596 ± 422, 359 ± 120 and 691 ± 445 %, respectively (**Figure 2G**). The significance of these measurements remained when the activities were normalised by VWF antigen (Ag) levels (**Figure 2H**) and a significant inverse relationship was observed between the ratio of VWF:CBA/Ag and ADAMTS13 concentration (**Figure 2J)**.

### Defective fibrin formation occurs in COVID-19 patient plasma

Fibrin formation was monitored *in vitro* using a turbidity-based assay (**Figures 3A and 3B)**. An increase in initial rate was observed in all COVID-19 sub-groups with a ~2-fold change in slope being seen in the moderate and severe groups (**Figure 3D**). Confocal imaging of fibrin clots, formed *in vitro*, was carried out to confirm that the increased initial rate of fibrin formation in patient plasma was the result of structural changes in the fibrin network (**Figure 3C**). In healthy control plasma (n=3) the fibrin network appeared to be evenly distributed, consisting of fine fibrin fibres with only small and few fibrin clusters (**Figure S2A**). In healthy control ‘clots’ there was very little VWF staining and it appeared to be largely localised within the small fibrin clusters (**Figure S2B**). A similarly even fibrin distribution was observed in all mild (n=3) and moderate (n=3) patient ‘clots’, albeit with larger and more abundant fibrin clusters. Plasma from patient 4 formed fibrin with some much larger fibrin clusters approaching 150 μm in length (**Figure S2A**). The most apparent change in fibrin structure was observed in the severe patients 18 and, in particular, patient 20 (**Figure S2A**). Again, VWF was co-localised with the large fibrin clusters and, in line with their increased size, was much more abundant (**Figure S2B**).

**Figure 3.**
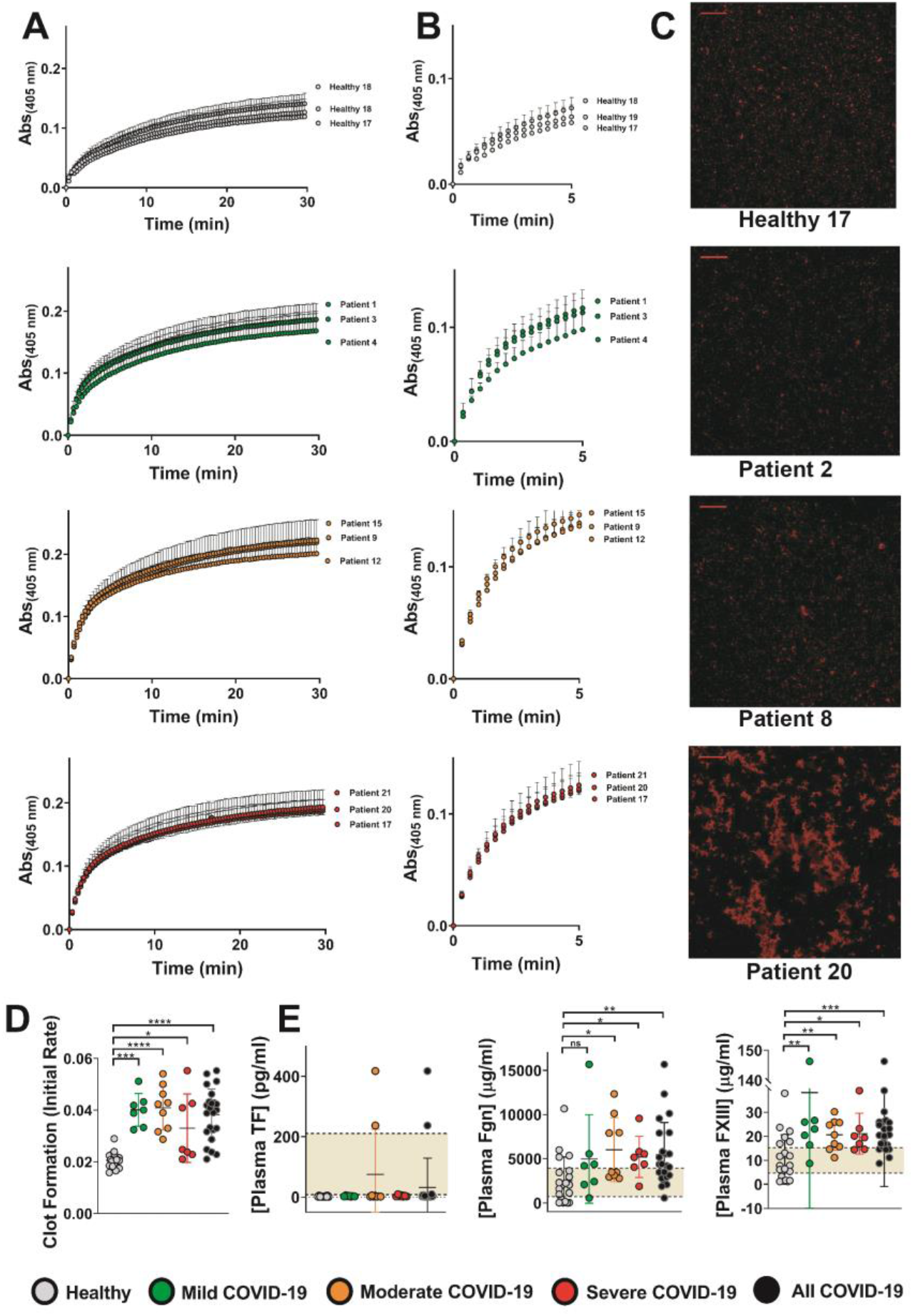
Thrombin-induced fibrin formation occurs at a faster rate in plasma from COVID-19 patients indicating a denser fibrin network. **(A)**, Turbidity assays were performed using plasma from healthy controls (n=20) and COVID-19 patient plasma (n=23). The absorbance measurements from 4 independent experiments (mean ± S.D.) are shown for 3 samples from each sub group (healthy, mild, moderate and severe COVID-19). **(B)**, the mean absorbance values for each sample over the first 3 minutes (9 measurements) were fitted to a linear regression model to determine the initial rate of change of fibrin formation (slope). **(C)**, the structure of the fibrin network, formed in the presence of AlexaFluor594 labelled fibrinogen (red), was visualised by confocal microscopy. Shown are example images from one sample in each subgroup. Scale bars represent 200 μm. Further imaging of a selection of samples from each group was also performed and can be found in Supplementary Figure 2. **(D)**, the initial rate values for fibrin formation in each sample were determined from 4 independent experiments and are presented as mean ± S.D. **(E)**, Plasma concentrations of tissue factor (TF), fibrinogen (Fgn) and Factor XIII (FXIII) were determined by LEGENDplex™ assay (human fibrinolysis 5-plex and human thrombosis custom 7-plex panels) as per manufacturer’s protocol. Each data point represents the mean of duplicate concentration values determined from fluorescence intensity and results are presented as mean ± S.D. Areas shaded in yellow represent the manufacturer’s normal range values (755-1698 μg/ml, 6.7-15.3 μg/ml and 5.9-200.9 pg/ml for Fgn, FXIII and TF respectively). Comparison of the COVID-19 population and healthy controls was performed using a Mann-Whitney test. Comparisons of COVID-19 sub-groups with healthy controls were performed with a Kruskal-Wallis test with Dunn’s correction for multiple comparisons (*<0.05, **p<0.01, ***p<0.001 and ****p<0.0001).

The concentration of tissue factor (TF) was within normal range (5.9-200.9 pg/ml) in all healthy controls and all COVID-19 patient samples, except 2 of the moderate cases (**Figure 3E**), with no significant difference in mean values between the groups. The concentration of fibrinogen in healthy controls was generally within either the manufacturer’s (0.75-1.7 mg/ml) or the clinical (2-4.5 mg/ml) normal range^24^ (**Figure 3E**). Of the 20 healthy controls only 4 were above the upper limit of normal, however, the mean value of 2.51 mg/ml was within range. There was a statistically significant increase in fibrinogen in the moderate and severe COVID-19 sub-groups, values all being above the upper limit of the normal range. A similar pattern of factor XIII (FXIII) concentrations was observed, with 14 out of the 20 healthy controls falling below the upper limit of the manufacturer’s normal range (6.7-15.3 μg/ml) and a mean value that was within this range (10.9 μg/ml). There was a significant increase in FXIII concentrations in all COVID-19 sub-groups (**Figure 3E**). Analysis of longitudinal samples from patient 21 identified a relationship between FXIII levels and impaired fibrin formation.

### Fibrin clot lysis by t-PA/plasmin is inhibited in COVID-19 patient plasma

The time to clot lysis in healthy control samples (**Figure 4A, 4C**) was consistently in the range of 15-20 minutes but was suppressed in COVID-19 patient plasma (**Figure 4B**), as demonstrated by the increase in mean lysis time across all severity groups (**Figure 4C)**. There appeared to be little effect of infection on the release of t-PA as the concentration in all healthy controls and most moderate and severe COVID-19 patient plasma was within, or marginally above, the normal range of 1.6-8.4 ng/ml (**Figure 4D**). Plasminogen levels were unaffected by COVID-19 infection and were all within the normal range of 79-170 μg/ml (**Table 1**). We observed a significant upregulation of PAI-1 in all COVID-19 sub-groups compared to healthy controls, of which, all but one were within the literature normal range of 20-100 ng/ml^25^, with COVID-19 samples all above this (**Figure 4D**). Analysis of longitudinal samples from patient 21 identified a relationship between PAI-1 concentration and impaired fibrinolysis. The concentrations of α2-antiplasmin were also influenced, with statistically significant increases observed in the mild and moderate COVID-19 sub-groups (**Figure 4E**). In healthy controls the plasma concentrations of α2-antiplasmin and TAFI were largely within the literature normal ranges of 50.4-85.4 μg/ml^16^ and 4-15 μg/ml^26^, respectively. The mean α2-antiplasmin concentrations in the mild, moderate and severe COVID-19 groups were all above the normal range, as was the mean TAFI concentration in the severe group (**Figure 4E**).

**Figure 4.**
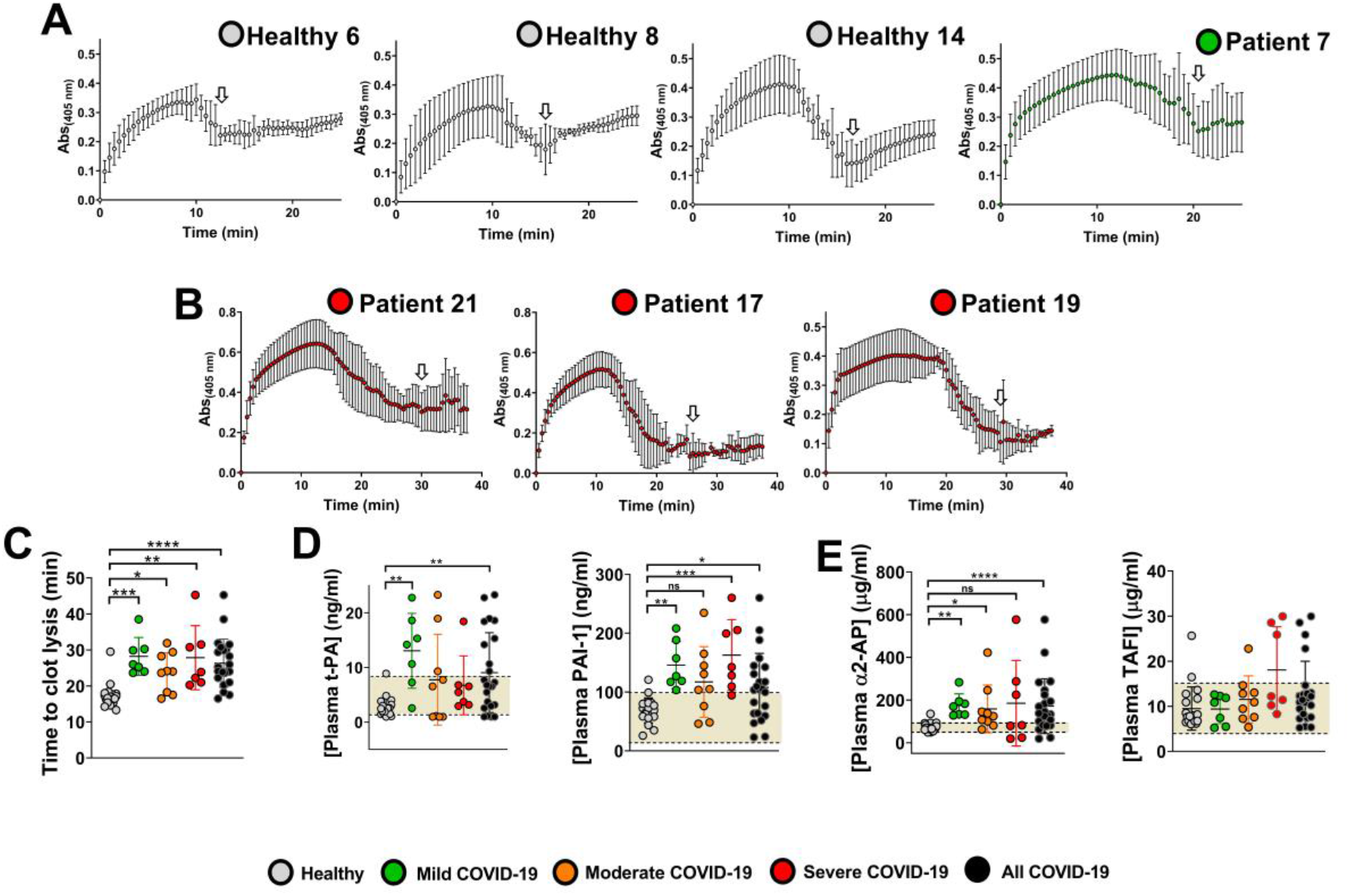
Fibrin formed in plasma from COVID-19 patients is more resistant to t-PA/plasmin-mediated fibrinolysis. Turbidity assays, in which fibrin formation is initiated by the addition of thrombin in the presence of t-PA, were performed using healthy control (n=20) and COVID-19 patient plasma (n=23). Shown are example traces from 3 healthy controls and a mild COVID-19 patient **(A)** and 3 severe COVID-19 patients **(B)**. For each, absorbance measurements are mean ± S.D. from 4 independent experiments. From each trace the mean time to clot lysis was manually determined (white arrows) and are presented for each sample as mean ± S.D. from the 4 replicates **(C)**. The plasma concentration of t-PA **(D)** was determined by LEGENDplex™ assay (human thrombosis custom 7-plex panel) and the concentrations of PAI-1 (**D**), α2-antiplasmin and TAFI (**E**) were determined by ELISA, as per manufacturer’s protocol. Each data point represents the mean of duplicate concentration values determined from PE fluorescence intensity or absorbance measurements, for LEGENDplex and ELISA respectively, and results are presented as mean ± S.D. Areas shaded in yellow represent the manufacturer’s normal range values (1.6-8.4 ng/ml for t-PA) or literature normal ranges (20-100 ng/ml, 50.4-85.4 μg/ml and 4-15 μg/ml for PAI-1, α2-antiplasmin and TAFI respectively). Comparison of the COVID-19 population and healthy controls was performed using a Mann-Whitney test. Comparisons of COVID-19 sub-groups with healthy controls were performed with a Kruskal-Wallis test with Dunn’s correction for multiple comparisons (*<0.05, **p<0.01, ***p<0.001 and ****p<0.0001).

**Figure 5.**
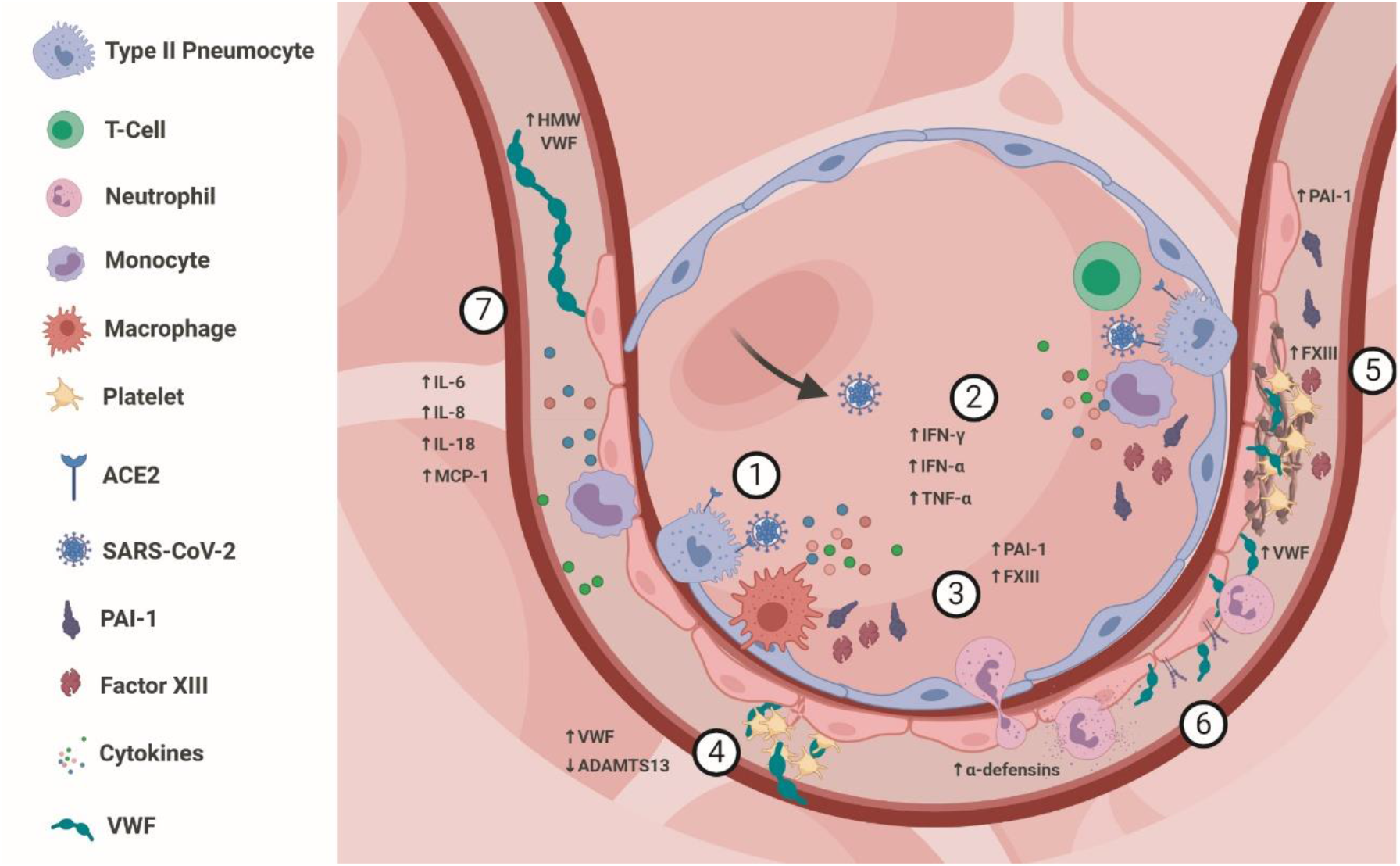
Proposed thromboinflammatory mechanisms underlying COVID-19 coagulopathy. **(1)**, SARS-CoV-2 infects Type II pneumocytes of the lung epithelium through binding to its ‘receptor’ angiotensin-converting enzyme (ACE)-2. **(2)**, infection elicits a type-1 interferon response (IFN-α) triggering the release of type-2 interferons (IFN-γ) and pro-inflammatory cytokines, including tumor necrosis factor (TNF)-a, from the epithelial cells and tissue resident leukocytes. **(3)**, resident macrophages, monocyte-derived macrophages and infiltrating monocytes express coagulation factor XIII (FXIII) and plasmin activator inhibitor (PAI)-1. **(4)**, endothelial activation induces Weibel-Palade body exocytosis and the release of von Willebrand factor (VWF). Concurrent depletion of ADAMTS13 results in the formation of ultra large (UL)-VWF multimers with high platelet reactivity and the formation of platelet aggregates. **(5)**, the activated endothelium further contributes to increased plasma PAI-1 and recruited platelets release FXIII. Together, these changes drive the formation of fibrinolytic resistant fibrin deposits. **(6)** up-regulation of cell adhesion molecules on the endothelium facilitates the further recruitment of leukocytes and propagation of the inflammatory response. The release of α-defensins from infiltrating neutrophils causes further endothelial activation and VWF/ADAMTS13 imbalance. **(7)**, together these immune and inflammatory responses result in a systemic pro-inflammatory state, through increased levels cytokines (interleukin (IL)-6, IL-8, IL-18 and monocyte chemoattractant protein (MCP)-1) and a systemic pro-coagulant state through increased circulating UL-VWF, FXIII, PAI-1 and α-defensins.

## Discussion

### A similar cytokine signature is present across all COVID-19 severity groups

The peripheral cytokine signature observed in this study is similar to that reported previously in a parallel study of the same patients^27^, with low levels of anti-viral cytokines (IFN-α and TNF-α) but substantial increases in pro-inflammatory cytokines (IL-6, IL-8, IL-10, IL-18, IL-23 and IL-33). This is true of all sub-groups, in particular the moderate and severe groups, in which the cytokine response is characteristic of ARDS of all causes^28^ and is reflected in a progressive increase in total white blood cell and neutrophil counts across the severity groups. Longitudinal analysis of the cytokine signature in severe patients is in agreement with a previous report suggesting the cytokine response diminishes in some patients as the disease progresses in severity and over time which may indicate exhaustion of some immune cell populations producing these cytokines^27^.

### COVID-19-associated coagulopathy is distinct from DIC

A screen of factors of the extrinsic, intrinsic and common coagulation cascades and key anti-coagulant pathways was performed in a smaller, initial cohort of samples and ruled out a role of these pathways in COVID-19 associated coagulopathy. Unlike in DIC, which was initially suspected as the cause of thrombotic complications in COVID-19 patients due to high D-dimer and PT^1^, there does not appear to be a consumption of coagulation factors, including Factor IX, which was quantified in the full cohort of samples. This is in agreement with the lack of bleeding phenotype observed in patients with COVID-19-associated coagulopathy^2^. While D-dimer was elevated significantly in the severe COVID-19 group, all values were within the expected normal range for the assay and the available clinical D-dimer measurements, although above the clinical normal range of <500 ng/ml, were far from the 5-20 μg/ml range that would suggest a diagnosis of DIC.

### VWF/ADAMTS13 imbalance is associated with increased IL-6 and neutrophil α-defensins

Increased plasma VWF has been suggested, in multiple recent studies^16-18^, to be a key driving force behind COVID-19 associated coagulopathy. This is further supported by the existence of VWF-platelet rich thrombi in the lungs of COVID-19 patients post-mortem which have been identified in numerous independent studies^29-31^. Our data is in agreement with this hypothesis as we report a substantial increase in plasma VWF, even in those with mild COVID-19 disease. It is likely that this increase in VWF is the result of endothelial activation and Weibel-Palade body exocytosis, which have been shown *in vitro* to be induced by IL-6^32^. As such, we observed a strong correlation between plasma VWF and IL-6.

We also report a substantial decrease in ADAMTS13 antigen in COVID-19 patients which, to date, has only been reported in one small case series^17^. As with VWF, this response is present in all COVID-19 sub-groups and correlates with IL-6. Although VWF and ADAMTS13 have both been implicated in other thrombotic pathologies, they are most often seen as independent risk factors^22, 23^. The fact that there is a strong correlation between VWF and ADAMTS13 in this cohort is unusual and may suggest that their increase and decrease, respectively, are driven by the same mediator, most likely IL-6. This is also indicated by longitudinal analysis, in particular that of patient 21, in which normal VWF/ADAMTS13 balance is restored as the IL-6 response diminishes. Other factors implicated in ADAMTS13 suppression are age (as with VWF), cytokine inhibition of hepatic synthesis^33^, direct inhibition by IL-6^32^ and direct inhibition by neutrophil α-defensins^34^. Interestingly, we report an increase in the plasma concentration of α-defensins in COVID-19 patients in our cohort and a robust inverse relationship between this and ADAMTS13 levels (R^2^=0.50, p<0.0001). Neutrophil counts and neutrophil activation (soluble PSGL-1) are both increased in these samples, in line with another recent study^35^, and both correlate with plasma VWF (R^2^=0.29, p<0.01 and R^2^=0.10, p<0.05, respectively).

The expected functional consequence of a concurrent increase in VWF and decrease in ADAMTS13 would be an increase in the multimeric size and therefore, activity, of VWF. Indeed, we report a significant increase in VWF collagen binding activity in all COVID-19 sub-groups with some moderate and severe patients exhibiting a >10-fold increase. This would be expected to have an impact on platelet-recruitment and might also enhance platelet aggregation and platelet-leukocyte aggregation which have been observed in recent studies^14, 36^. Platelet counts in healthy controls were similar to that of patients therefore no relationship between increased VWF and platelet depletion could be identified in this cohort of COVID-19 patients. Mild thrombocytopenia has been reported previously in COVID-19 patients^37^ but is only evident in three patients in this cohort and there is no apparent relationship between platelet count and VWF. Interestingly, in patient 21, in which VWF levels normalise in later samples, there is a corresponding increase in platelet count into the normal range.

### Platelet-derived FXIII alters *in vitro* fibrin formation and clot structure

Perhaps the most universally altered parameter amongst COVID patient plasma is an increase in initial rate of *in vitro* fibrin formation. This does not appear to be the result of increased fibrinogen levels, observed here and in previous reports^38, 39^, as there is little variation in initial rate within the healthy control group despite differing fibrinogen concentrations. There is also a relatively weak correlation between fibrinogen concentration and initial rate (R^2^=0.19, p=0.003). Instead, the alteration is likely the result of increased incorporation of VWF into the fibrin network^40, 41^ and FXIII-mediated fibrin fibre compaction^42^. This is supported by a significant correlation between initial rate and both VWF (R^2=^0.45, p<0.0001) and FXIII (R^2^=0.30, p=0.0002) and by a longitudinal relationship between FXIII (increasing to above the normal range) and increased initial rate in patient 21. Furthermore, defective fibrin structure, identified by confocal imaging, is more apparent in patients who have FXIII levels above the normal range and particularly high VWF levels.

In those patients, large fibrin clusters were formed which contained VWF, mirroring the composition of thrombi identified in post-mortem lung examinations in COVID-19 patients^30^. These structures may derive from the fact that FXIII-mediated crosslinking of fibrin results in a higher density of fibres^20^ and that FXIII is known to covalently crosslink VWF to fibrin^43^. Although the precise nature of this fibrin structure defect, and its potential impact on clot permeability, are not yet fully understood it is possible that the large fibrin clusters formed may be impermeable which might be expected to influence fibrinolysis (see below). Likely sources of the increased levels of FXIII in COVID-19 patients are monocytes, macrophages and platelets recruited to the pulmonary endothelium. Given the relatively normal number of circulating monocytes in all COVID-19 sub-groups, platelets appear to be the likely source of FXIII. Increased numbers of activated platelets have been identified as a common feature amongst COVID-19 patients^14, 36^ and is indicated in this cohort by increased plasma P-selectin and soluble CD40L.

### Altered clot structure and endothelium-derived PAI-1 induce fibrinolytic resistance

Based on the defective fibrin formation observed in COVID-19 patient plasma herein, and the previous identification of persistent fibrin deposits and hyaline membranes in post-mortem lung samples^30^, we hypothesised that fibrin formed in the pulmonary vasculature and alveolar spaces may be resistant to endogenous fibrinolysis. Using an *in vitro* assay of t-PA/plasmin-mediated fibrinolysis we have identified a ~2 fold extension of clot lysis times, irrespective of disease severity. This may be influenced, in part, by the altered fibrin structure described above and this is supported by a correlation between the initial rate of fibrin formation and time to clot lysis (R^2^=0.75, p<0.0001).

There are four main inhibitory factors that supress fibrinolysis by the t-PA/plasmin pathway, PAI-1, α2-antiplasmin, α2-macroglobulin and TAFI. Although they are all primarily produced by the liver, their plasma concentrations can be potentially influenced by inflammatory and immune responses known to be involved in COVID-19. The factor most significantly affected by COVID-19 seems to be PAI-1, with almost all patients exhibiting increased levels despite relatively normal levels of t-PA. The importance of this imbalance is reflected in the correlation between PAI-1 and an increased time to clot lysis (R^2^=0.22, p<0.002). This is unsurprising as PAI-1 is known to be released from the activated endothelium, reported in a previous COVID-19 cohort^16^ and evident in this cohort by increased plasma VWF, FVIII and P-selectin. Given the size of the vascular bed in the lungs this is likely to account for the majority of the PAI-1 increase, although there may also be a contribution from monocytes and macrophages in the pulmonary vasculature. Increased PAI-1 has been linked to obesity and identified in non-diabetic obese patients (BMI >30)^44^, however, most participants in this study had a BMI <30 and this is unlikely to account for the large observed increase.

The impact of COVID-19 on the levels of α2-antiplasmin and TAFI, released locally at low levels by activated platelets, is arguably less significant. In the case of α2-antiplasmin there is a reasonably strong correlation between α2-antiplasmin and time to clot lysis (R^2^=0.27, p=0.0003) indicating a disproportionate inhibitory effect. This is likely to be due to an enhancement of α2-antiplasmin’s inhibitory potential caused by the concurrent increase in FXIII, thus increasing its incorporation into the fibrin network as it forms.

## Summary

It should be noted that the haemostatic alterations identified in this cohort, and thrombotic complications of COVID19 reported previously^5^, have occurred despite most patients receiving thromboprophylaxis and, in some cases, treatment doses of anti-coagulants. Therefore, this may have implications in the ongoing management of such complications in patients who may require higher prophylactic doses while hospitalised and extended anticoagulation post-discharge. Until we have a complete understanding of the mechanism involved it may be difficult to predict which common anti-coagulant therapy, if any, is most appropriate. Given the apparent role of FXIII and VWF in the defective coagulation, experimental therapies targeting these factors such as the FXIII inhibitor tridegin^45^ and rADAMTS13^46, 47^, are potentially worth pursuing. The apparent fibrinolytic resistance in COVID-19 patients may also impact on the efficacy of rt-PA thrombolysis to treat hyperacute ischaemic stroke or massive pulmonary embolism complicating SARS-CoV-2 infection and, again, experimental avenues might need to be explored to overcome this. Potential new thrombolytic therapies such as rADAMTS13, which is unlikely to be influenced by fibrinolytic inhibition, or α2-antiplasmin inhibitors, with the ability to correct the defective fibrinolysis thereby restoring the efficacy of rt-PA thrombolysis^48^, are both viable options that should be explored.

## Data Availability

Data available on request.

## Acknowledgements

This work was funded by Medical Research Foundation Fellowship (MRF-076-0004-RG-SOUT-C0756) awarded to K. South. This report is independent research supported by the North West Lung Centre Charity and the National Institute for Health Research Manchester Clinical Research Facility at Manchester University NHS Foundation Trust (Wythenshawe). We acknowledge the Manchester Allergy, Respiratory and Thoracic Surgery Biobank for supporting this project and thank the study participants for their contribution. Angela Simpson, Alexander Horsely and Tim Felton are supported by the Manchester Biomedical Research Centre. Imaging was performed in the University of Manchester Bioimaging core facility. Flow cytometry and LEGENDplex assays were performed in the University of Manchester Flow Cytometry core facility.

## Authors

### CIRCO investigators

Rohan Ahmed, Halima Ali Shuwa, Miriam Avery, Katharine Birchall, Oliver Brand, Evelyn Charsley, Alistair Chenery, Christine Chew, Richard Clark, Emma Connolly, Karen Connolly, Paul Dark, Simon Dawson, Laura Durrans, Hannah Durrington, Jasmine Egan, Claire Fox, Helen Francis, Miriam Franklin, Susannah Glasgow, Nicola Godfrey, Kathryn J. Gray, Seamus Grundy, Jacinta Guerin, Pamela Hackney, Mudassar Iqbal, Chantelle Hayes, Emma Hardy, Jade Harris, Anu John, Bethany Jolly, Verena Kästele, Saba Khan, Gabriella Lindergard, Graham Lord, Sylvia Lui, Lesley Lowe, Alex G Mathioudakis, Flora A. McClure, Joanne Mitchell, Clare Moizer, Katrina Moore, David J. Morgan, Stuart Moss, Syed Murtuza Baker, Rob Oliver, Grace Padden, Christina Parkinson, Laurence Pearmain, Mike Phuychareon, Ananya Saha, Barbora Salcman, Nicholas A. Scott, Seema Sharma, Jane Shaw, Joanne Shaw, Elizabeth Shepley, Lara Smith, Simon Stephan, Ruth Stephens, Gael Tavernier, Rhys Tudge, Louis Wareing, Roanna Warren, Thomas Williams, Lisa Willmore, Mehwish Younas,

## Authorship Contributions

K.S., performed the data collection and generated the figures and K.S., L.M., L.R. and J.G. analysed the data.

K.S., J.G., T.H., C.S. and S.A. designed the study, interpreted the data and wrote the manuscript.

E.M., M.M. and S.K. performed sample collection and biobanking.

PD, AS, TF, AH identified the participants and supervised recruitment and designed the clinical follow up.

## Disclosure of Conflicts of Interest

The authors declare no conflicts of interest

Supplementary Material

**Figure S1.**
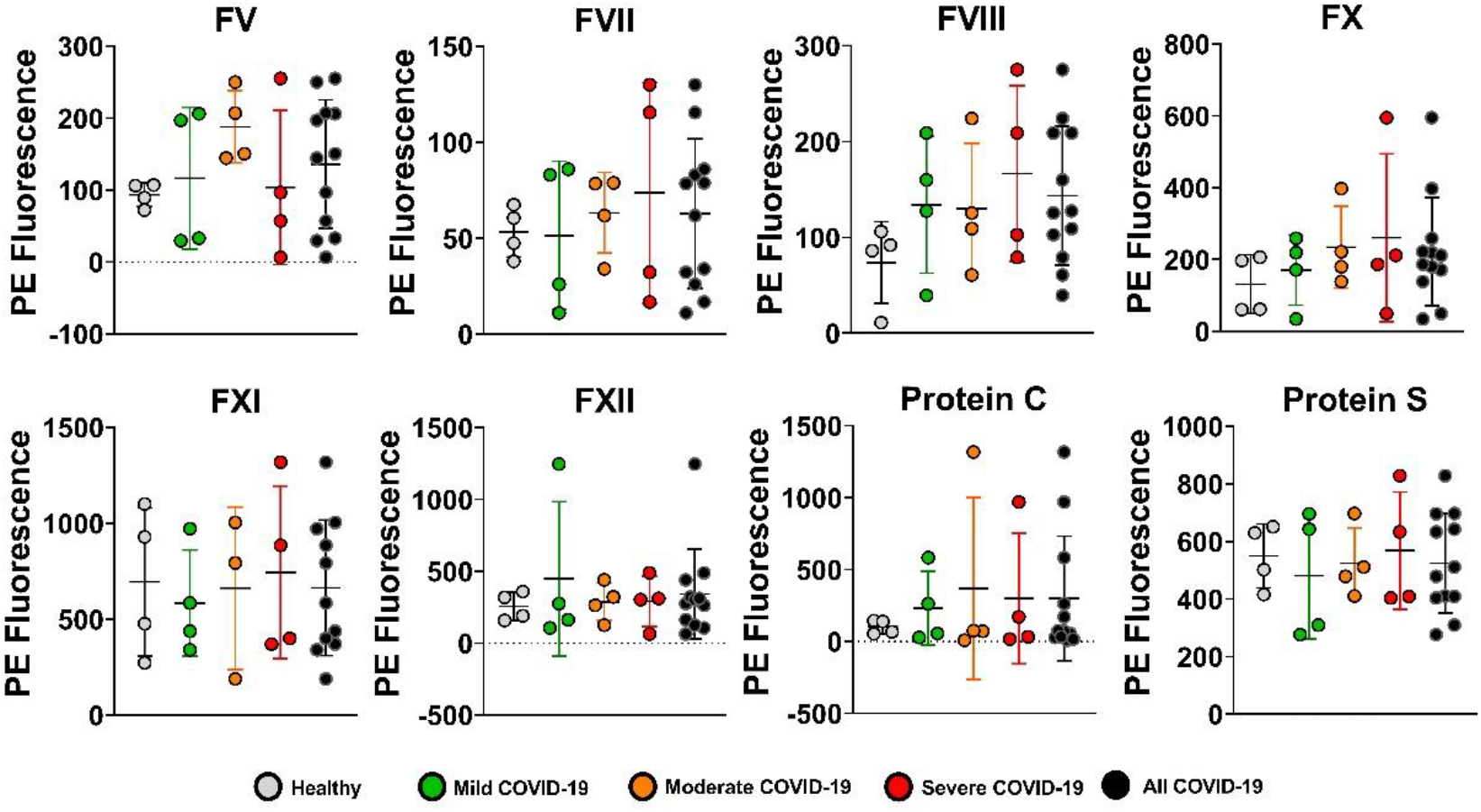
The concentrations of many plasma coagulation factors are not significantly altered in COVID-19 patients. The plasma concentrations of coagulation factors were determined by ProcartaPlex™ assay (coagulation panels 1-3) as per manufacturer’s protocol. Each data point represents the mean of duplicate PE fluorescence measurements. Results are presented as mean ± S.D. for both healthy control (n=4) and COVID-19 patient plasma (n=11-12). In the moderate group a sampling error reduced the number of FXI readings to n=3. Comparison of the COVID-19 population and healthy controls was performed using a Mann-Whitney test and showed no significant difference.

**Figure S2.**
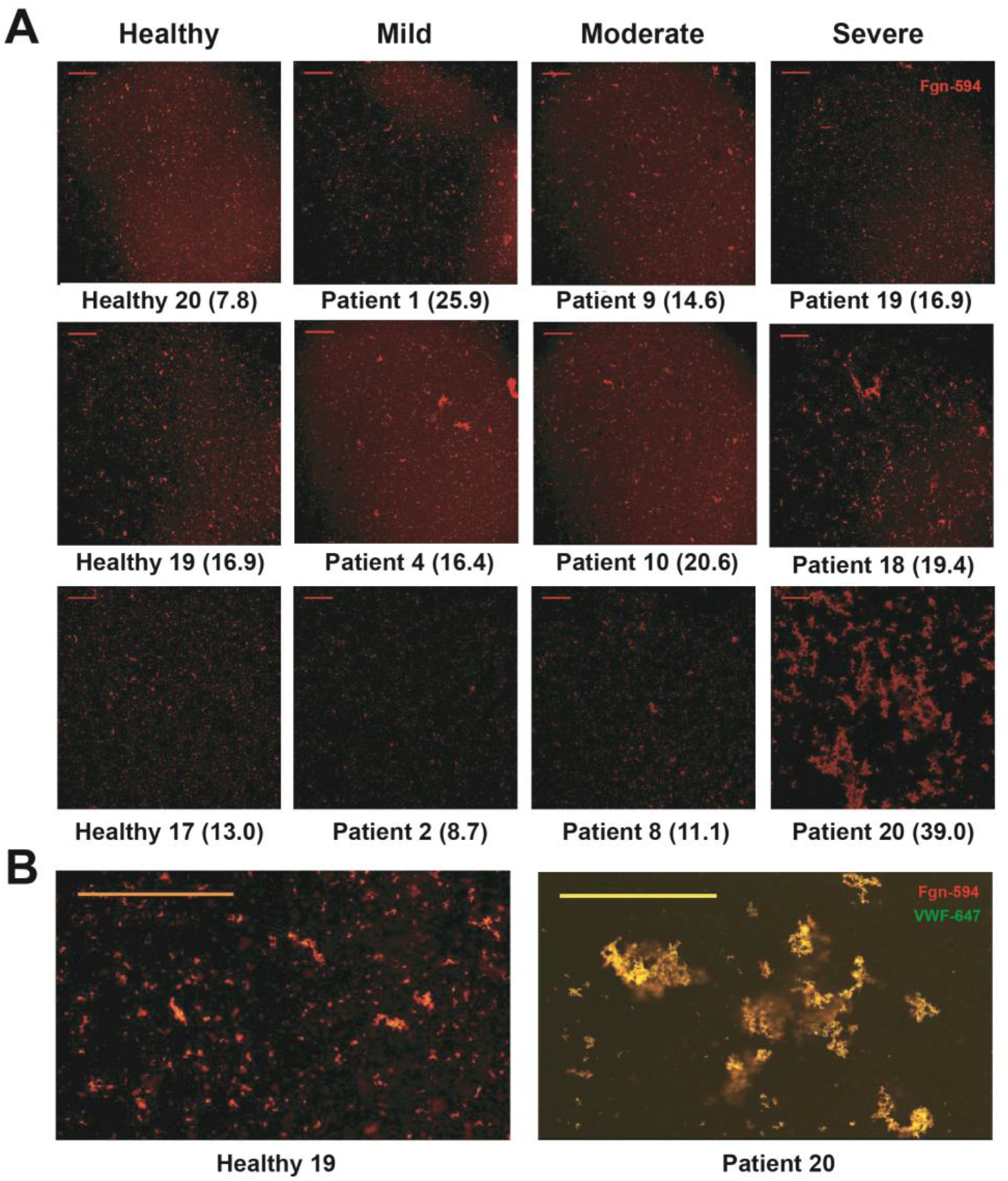
The microscopic structure of fibrin in *in vitro* ‘clots’. **A**, fibrin clots, formed in plasma from 3 healthy controls and 3 patients from each COVID-19 sub-group, were imaged by confocal microscopy. Selection of samples from COVID-19 groups was based on FXIII concentrations (numbers in brackets) which were either within, or above, the normal range of 6.7-15.3 μg/ml. **B**, example images at higher magnification showing co-localisation (yellow) of fibrin clusters and VWF staining. Scale bars represent 200 μm.

**Figure S3.**
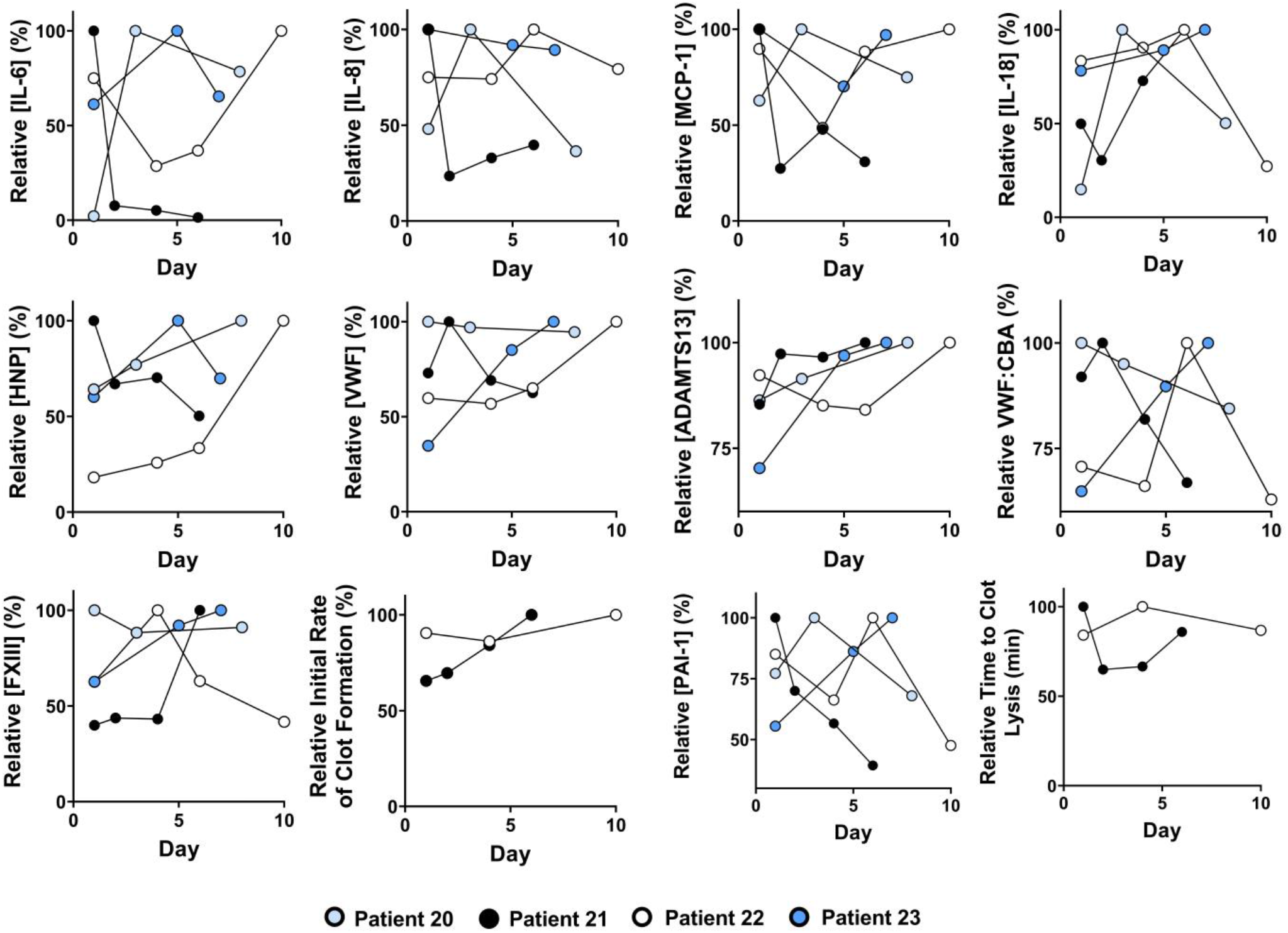
Longitudinal analysis of thromboinflammatory parameters in severe COVID-19 patients. Plasma at each time point was immediately banked upon collection and all samples were analysed at the same time and alongside all other healthy and patient samples. The x-axis refers to the number of days elapsed since ICU admission. Data for each individual parameter has been normalised for each sample relative to the maximal response observed in that patient series. Due to the low volume of plasma available for some time points in patients 20 and 23, fibrin formation and fibrinolysis assays were not performed.

